# Epidemiological insight into the possible drivers of Lassa fever in an endemic area of Southwestern Nigeria from 2017 and 2021

**DOI:** 10.1101/2023.09.06.23295111

**Authors:** Simeon Cadmus, Victor Akinseye, Eniola Cadmus, Gboyega Famokun, Stephen Fagbemi, Gabriel Ogunde, Ayuba Philip, Rashid Ansumana, Adekunle Ayinmode, Taiwo Olalekan, Oladimeji Oluwayelu, Oyewale Tomori, Solomon O. Odemuyiwa

## Abstract

**Background:** Lassa fever (LF) is a viral disease transmitted between animals and humans, commonly found in West Africa, including Nigeria. The region experiences an estimated annual total of about 2 million LF cases in humans, leading to 5,000 to 10,000 deaths. Strikingly, up to 80% of LF-infected individuals show no symptoms, making its true incidence hard to determine in endemic populations. We investigated LF distribution, mortality, survival patterns, and contributing factors during a local outbreak in Nigeria, from 2017 to 2021.

**Method:** Data from the Integrated Disease Surveillance and Response weekly line list for 2017 to 2021 were extracted. The survival pattern of LF patients was visualized with the Kaplan-Meier curve, binary logistic regression model was employed to explore LF-associated factors and level of statistical significance (α) was set at 5%.

**Result:** Overall, 4,554 participants were recruited between 2017 and 2021. Their average age varied from 31.82 ± 20.0 to 37.85 ± 17.89. LF-positive patients decreased from 26.9% in 2017 to 17.7% in 2021, paralleling the mortality trend. In 2021, patient survival ranged from 5 to 30 days. Male patients had lower survival odds in the initial 10 days of hospitalization, improved chances from days 10 to 20, and reduced probabilities beyond day 20. Residence location and age were significant factors (p<0.05) associated with LF in Ondo State.

**Conclusion:** The decline in LF cases in 2021 could be attributed to the ongoing intervention by Nigerian Centre for Disease Control or the disruption caused by the COVID-19 pandemic in 2020. To address LF challenges in hotspot areas, we propose Community Action Networks that would operate using the One Health approach involving local stakeholders sustainably to promote Early Warning/Early Response system in high-risk settings and mitigate LF-related issues.

**SUMMARY:** Lassa fever (LF) is an important disease of global public health concern that is endemic in West Africa. In Nigeria, the disease constitutes a major health challenge with outbreaks being recorded on an annual basis despite efforts channeled towards combating it by the government at various levels. This study analysed a five years data of LF in Ondo State southwestern Nigeria. The results identified age and location were identified as important factors associated with infection and mortality among LF patients as the incidence and case fatality rates were highest among adults (≥ 45 years), while the highest number of suspected, confirmed and dead cases was recorded in Owo Local Government Area. Furthermore, we identified drying of food items by the roadside where rodent vectors can access them, presence of a local market, poor and unsafe sewage disposal, and proximity of refuse dumps to residential areas as possible socio-ecological factors/practices fueling the endemicity and seasonal outbreak of LF. These findings emphasize the need for active involvement of community members in the already established national LF surveillance network to facilitate prompt case identification, and early reporting and response in the LF-endemic areas of the country.

## BACKGROUND

Lassa fever (LF) is a neglected viral zoonosis endemic in Nigeria, Benin Republic, and the Mano River Union countries (Sierra Leone, Guinea, Liberia) of West Africa [1, 2]. Sporadic outbreaks of LF have also been reported in Ghana [5], Mali, Senegal, and Cote d’Ivoire [6] [6, 11–13] (Figure 1). In this region, it is estimated that approximately 2 million human infections and 5,000-10,000 deaths are associated with LF annually [7]. In the healthcare environment, the disease is transmitted from person to person through contact with infected individuals as a result of poor biosafety practices [7]. Conversely, in the general population, disease transmission is indirect and associated with contamination of food with feces and urine from the reproductively prolific multimmamate rat, *Mastomysis natalensis* [8]. Thus, in endemic countries like Nigeria, LF outbreaks are common in rural areas where agricultural practices and sociodemographic factors favor human-rodent interactions.

**Figure 1:**
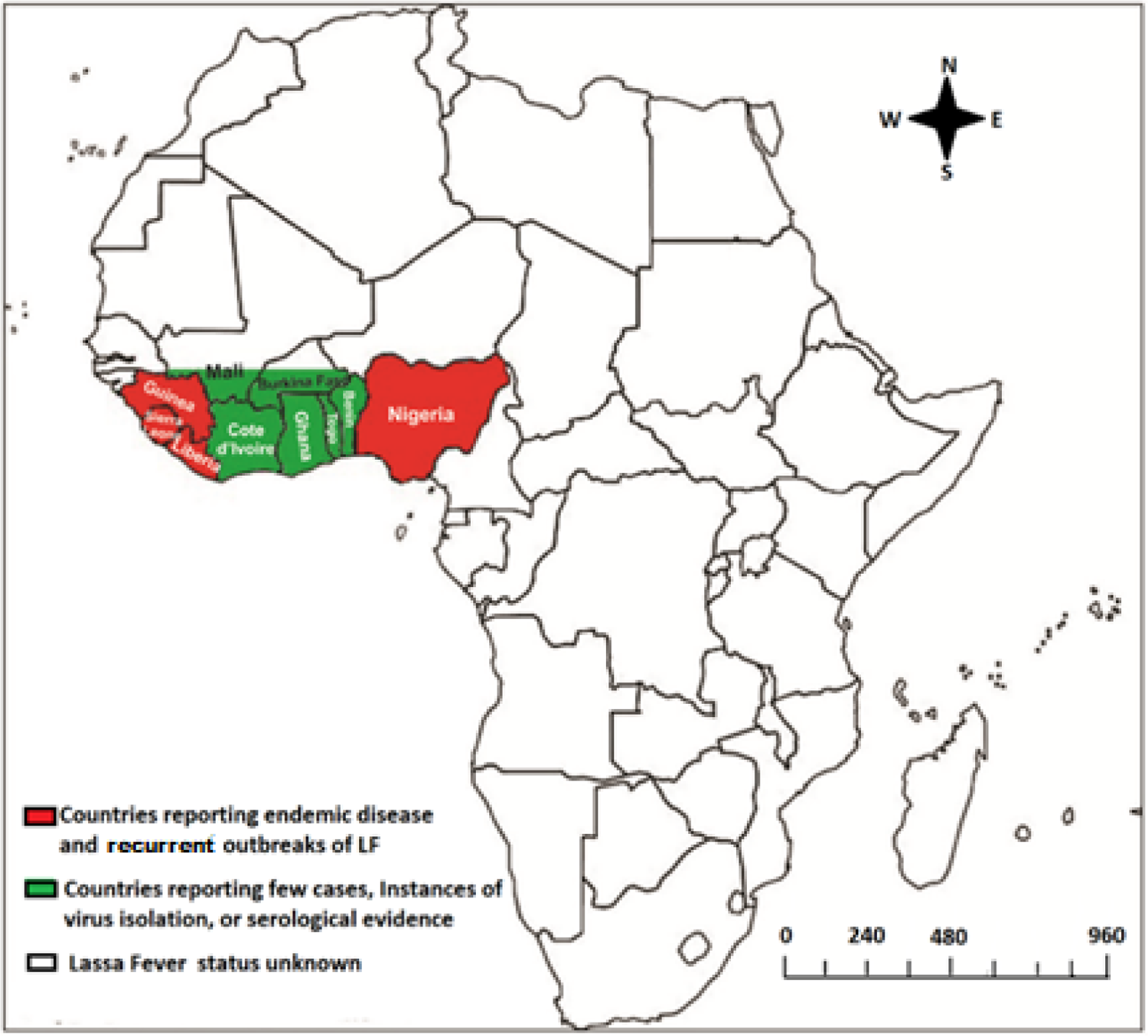
Map of Africa showing geographic distribution of Lassa fever. NB: Data for this map was obtained from CDC, 2022

LF is caused by Lassa fever virus (LASV), a single-stranded, segmented, RNA virus belonging to the Arenaviridae family [1, 8]. Phylogeographic analyses have classified LASV into six main clades that appear to be associated with specific geographic areas in West Africa. Isolates belonging to clades I - III are found in Nigeria, clade IV exists in Sierra Leone, Guinea, and Liberia, clade V is described in southern Mali, while recent isolates from Togo were classified as clade VI. Although the seeming geographic restriction of LFV genotypes suggests adaptation to local reservoir hosts, like most RNA viruses, continuous evolution of isolates within their natural nests will ensure the emergence of new phenotypic and genotypic variants over time [9, 10]. Such changes may result in different clinical presentation of LF and affect the course and outcomes of infection. It is therefore important to evaluate changes in the course of disease in endemic areas to identify patterns that may reflect altered pathogenicity of local genetic variants of LFV.

The incubation period of LF has been estimated as 3-21 days following LFV infection. However, up to 80% of individuals infected with LFV are asymptomatic or show mild non-differentiating clinical signs and symptoms that may not require treatment. Therefore, the disease may remain undetected in a community for a long time until severe cases occur, making it difficult to determine the true incidence of LF in an endemic population [9, 11–13]. In this report, we analyzed mortality and survival patterns during a local outbreak of LF in Ondo state, Nigeria between 2017 and 2022. We evaluated sociodemographic factors associated with LFV infection and survival during an outbreak.

## METHODS

### Study context

Between January 1 and December 31, 2022, a total of 8,201 suspected cases of LF were reported in Nigeria with 1,067 (13.0%) confirmed and 189 deaths (CFR = 17.7%). Overall, 112 Local Government Areas (LGAs) across 27 States of Nigeria reported at least one confirmed case over this period in 2022. Nonetheless, three states accounted for about 72% of the cases reported (Ondo: 33%, Edo: 25% and Bauchi: 14%) [14]. Thus, Ondo State, for the first time since 2017, has now overtaken Edo State as the epicenter of LF in Nigeria [15]. This sudden upsurge in the number of LF cases in Ondo State presents an opportunity to examine demographic factors associated with mortality during an outbreak of LFV in an endemic area. We therefore explored LF surveillance data for Ondo State from 2017-2021 with the aim of gaining better insight into the trends and possible predictors of the course of the disease in the state.

### Geographic location

Ondo State is one of the 36 states of the Federal Republic of Nigeria. It is situated in the southwestern region of the country. The state lies between longitudes 4“30” and 6” East, and latitudes 5“45” to 8” 15” North. It has land borders with Ekiti and Kogi State to the North, Edo State to the East, Oyo and Ogun State to the West, and the Atlantic Ocean to the South. Ondo State has a total land area of 14,788,723sq km. and a projected population of 4,883,793 (male =2,462,525; female = 2,421,267). It is divided into 18 LGAs, namely, Akoko Northeast, Akoko Northwest, Akoko Southeast, Akoko Southwest, Akure North, Akure South, Ose Odo, Idanre, Ifedore, Ilaje, Ileoluji/Okeigbo, Irele, Odigbo, Okitipupa, Ondo East, Ondo West, Owo, and Ose. The state’s ethnic composition includes members of the Akoko, Akure, Apoi, Idanre, Ijaw, Ikale, Ilaje, Ondo, and Owo groups. The region has a typical tropical climate with two main seasons: the rainy season (April to October) and the dry season (November to March). Throughout the year, temperatures fluctuate between 21°C and 29°C, and humidity is very high. The annual rainfall is estimated at 1,150-2,000 mm. There is a lot of arable soil that is good for farming. The northern edges have sub-savannah forests suitable for grazing cattle. The state has a vast forest that contains a variety of timber species. Ondo State is the largest producer of cocoa in Nigeria. Rubber, cashews, kolanuts, palm oil, and other economic crops are also farmed in the state. The State contributes approximately 12% of Nigeria’s overall oil and gas production [16].

### Data source

Data was obtained from the Epidemiology Unit of Ondo State Ministry of Health. Only cases that had met the Nigeria Centre for Disease Control (NCDC) case definition for LF and had laboratory results entered were considered for the study. Cases with pending laboratory results were excluded. The data analyzed in this report were extracted from the Integrated Disease Surveillance and Response (IDSR) weekly epidemiological data line list for 2017 to 2021. Curated data were exported and saved as excel files.

### Definition of study variables

Independent variables: The major independent variables selected were LGA, date of onset of symptoms, date seen at the health facility, time to presentation at the hospital, in-patient (Yes/No), sex, and age of patient. Dependent/outcome variables: The outcome variables include result of laboratory confirmatory testing for LFV infection (positive/negative) and outcome (alive, dead). Others include year and month of disease occurrence.

### IDSR Lassa fever case definitions

**A suspected case**: any person with an illness of gradual onset with one or more of the following: malaise, fever, headache, sore throat, cough, nausea, vomiting, diarrhoea, myalgia (muscle pain), central chest pain or retrosternal pain, hearing loss and either a history of contact with excreta or urine of rodents or history of contact with a probable or confirmed LF case within a period of 21 days of onset of symptoms or any person with inexplicable bleeding.

**A probable case:** any suspected case as defined above but who died or absconded without collection of specimens for laboratory testing.

**A confirmed case**: any suspected case with laboratory confirmation (positive IgM antibody, PCR or virus isolation).

**Ethical considerations:** approval was received from the Surveillance and Epidemiology Unit of Ondo State Ministry of Health and University of Ibadan/University College Hospital Institutional Review Board (UI/UCH/22/0305).

**Data analysis**: Data for the period under review were sorted, cleaned and relevant variables were extracted using Microsoft Excel. Cleaned data were imported into EpiInfo® Software version 7.0. Descriptive statistics (mean, SD, frequency and proportions) was used to appropriately describe the dataset. ArcGIS was used to fine tune the geographical clustering of LF in Ondo State. The Kaplan Meier curve was employed to describe the survival pattern of LF patients. Finally, a binary logistic regression model was used to investigate the factors that were associated with LF. The level of statistical significance (α) was set at 5%.

## RESULTS

### Socio-demographic Pattern of Lassa Fever Patients in Ondo State (2017-2021)

The present study involved a comprehensive analysis of LF patients over a five-year period, encompassing data from 2017 to 2021. The study included a progressively increasing number of participants each year, starting with 283 participants in 2017, 626 participants in 2018, 1,123 in 2019, 1,558 in 2020, and 964 participants in 2021 (Table 1). The participant number reflected the growing scope and scale of the study over time, allowing for more robust and representative findings.

**Table 1:** Socio-demographic Pattern of Lassa fever Patients in Ondo State (2017-2021)

|  | 2017 (n=283)<br>n (%) | 2018 (n=626)<br>n (%) | 2019 (n=1123)<br>n (%) | 2020 (n=1558)<br>n (%) | 2021 (n=964)<br>n (%) |
| --- | --- | --- | --- | --- | --- |
| **Result** |  |  |  |  |  |
| Negative | 207 (73.1) | 463 (74.0) | 833 (74.2) | 1132 (72.7) | 791 (82.1) |
| Positive | 76 (26.9) | 162 (25.8) | 282 (25.1) | 424 (27.2) | 171 (17.7) |
| NA | 0 | 1 (0.2) | 8 (0.7) | 2 (0.1) | 2 (0.2) |
| **Mean age ± SD** | 37.85 ±<br>17.89 | 35.06 ±<br>20.13 | 31.82 ± 20.0 | 34.12 ±<br>21.29 | 37.33 ±<br>21.97 |
| **Age group (yrs)** |  |  |  |  |  |
| < 20 | 37 (13.1) | 137 (22.1) | 306 (27.3) | 382 (24.5) | 219 (22.7) |
| 20 – 29 | 60 (21.2) | 122 (19.7) | 227 (20.2) | 322 (20.7) | 159 (16.5) |
| 30 – 39 | 60 (21.2) | 138 (22.3) | 237 (21.1) | 236 (15.2) | 161 (16.7) |
| 40 – 49 | 56 (19.8) | 90 (14.5) | 139 (12.4) | 200 (12.8) | 125 (13.0) |
| 50 – 59 | 19 (6.7) | 53 (8.6) | 82 (7.3) | 143 (9.2) | 95 (9.8) |
| 60+ | 42 (14.8) | 80 (12.9) | 115 (10.2) | 224 (14.4) | 180 (18.7) |
| Non-response | 9 (3.2) | 6 (1.0) | 17 (1.5) | 51 (3.3) | 25 (2.6) |
| **LGA** |  |  |  |  |  |
| Akoko | 18 (6.4) | 32 (5.1) | 86 (7.7) | 177 (11.4) | 78 (8.1) |
| Akure | 41 (14.5) | 103 (16.5) | 189 (16.8) | 327 (21.0) | 189 (19.6) |
| Ose | 33 (11.7) | 47 (7.5) | 110 (9.8) | 101 (6.5) | 61 (6.3) |
| Owo | 182 (64.3) | 392 (62.6) | 695 (61.9) | 900 (57.8) | 600 (62.2) |
| Others | 9 (3.2) | 52 (8.3) | 43 (3.8) | 53 (3.4) | 36 (3.7) |
| **Sex** |  |  |  |  |  |
| Female | 126 (44.5) | 291 (46.5) | 558 (49.8) | 794 (51.0) | 471 (49.0) |
| Male | 155 (54.8) | 332 (53.0) | 563 (50.2) | 753 (48.3) | 490 (51.0) |
| NA | 2 (0.7) | 3 (0.5) | 2 (0.2) | 11 (0.7) | 3 (0.3) |
| **In-Patient** |  |  |  |  |  |
| No | 5 (1.8) | 4 (0.6) | 1 (0.1) | 0 | 0 |
| Yes | 278 (98.2) | 621 (99.2) | 1122 (99.9) | 1558 (100.0) | 964 (100.0) |
| Unknown | 0 | 1 (0.2) | 0 | 0 | 0 |
| **Outcome** |  |  |  |  |  |
| Alive | 260 (91.9) | 575 (92.4) | 1049 (93.4) | 1469 (94.3) | 913 (94.7) |
| Dead | 23 (8.1) | 47 (7.6) | 74 (6.6) | 89 (5.7) | 51 (5.3) |
| Unknown | 0 | 4 (0.6) | 0 | 0 | 0 |

Among the 283 participants recruited in 2017, the mean age was estimated to be 37.85 ± 17.89 years. This average age remained consistent throughout the subsequent years, indicating that the participants’ age profile was similar across the study period. The consistency in age distribution ensured that any observed changes in LF prevalence or mortality rates were less likely to be influenced by age-related factors.

In 2017, out of the 283 participants, 76 individuals were identified as LF-positive patients, constituting 26.9% of the cohort. However, in the following years, the proportion of positive cases gradually decreased. By 2021, only 17.7% of the participants were confirmed as LF cases. Notably, in 2020, there was a temporary peak in LF cases, with 27.2% of participants testing positive (Table 1). Throughout the five-year study period, majority (approximately 60%) of the participants lived in Owo LGA. Furthermore, there was a decline in the percentage mortality among participants during the period. In 2017, the mortality rate was 8.1%, which gradually decreased to 5.3% in 2021 (Table 1).

### Distribution of suspected and confirmed cases, and mortality among LF cases, Ondo State, 2017-2021

A total of 4,546 suspected cases were reported over the period under review. Of these, 1,113 (24.5%) were positive and 12 (0.3%) probable. There were 284 fatalities (case fatality rate, CFR, 25.5%). Males were generally more affected than females [578 (25.2%) positive; 161 deaths, CFR=27.9%)] (Table 2). The highest number of cases were reported in Owo (60.8%) and Akure South (13.7%), two LGAs that are in close (Figure 2; Figure 3). Slightly more deaths (n=143) were recorded among patients who reported to the hospital after 7 days of onset of symptoms compared to those who reported earlier (n=141) (Table 2).

**Figure 2:**
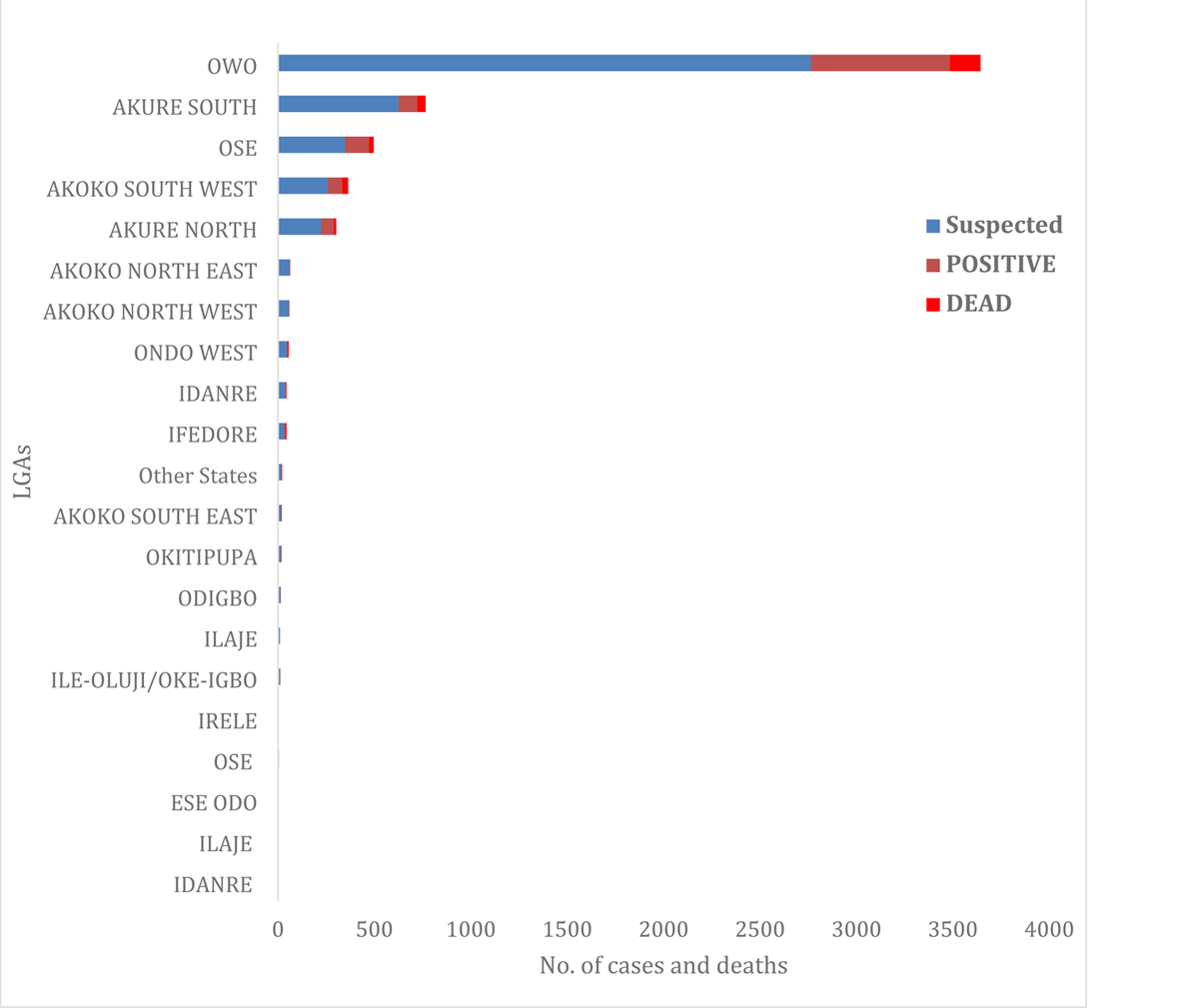
The distribution of suspected and confirmed cases and deaths by LGA based on Ondo State LF data, 2017-2021.

**Figure 3:**
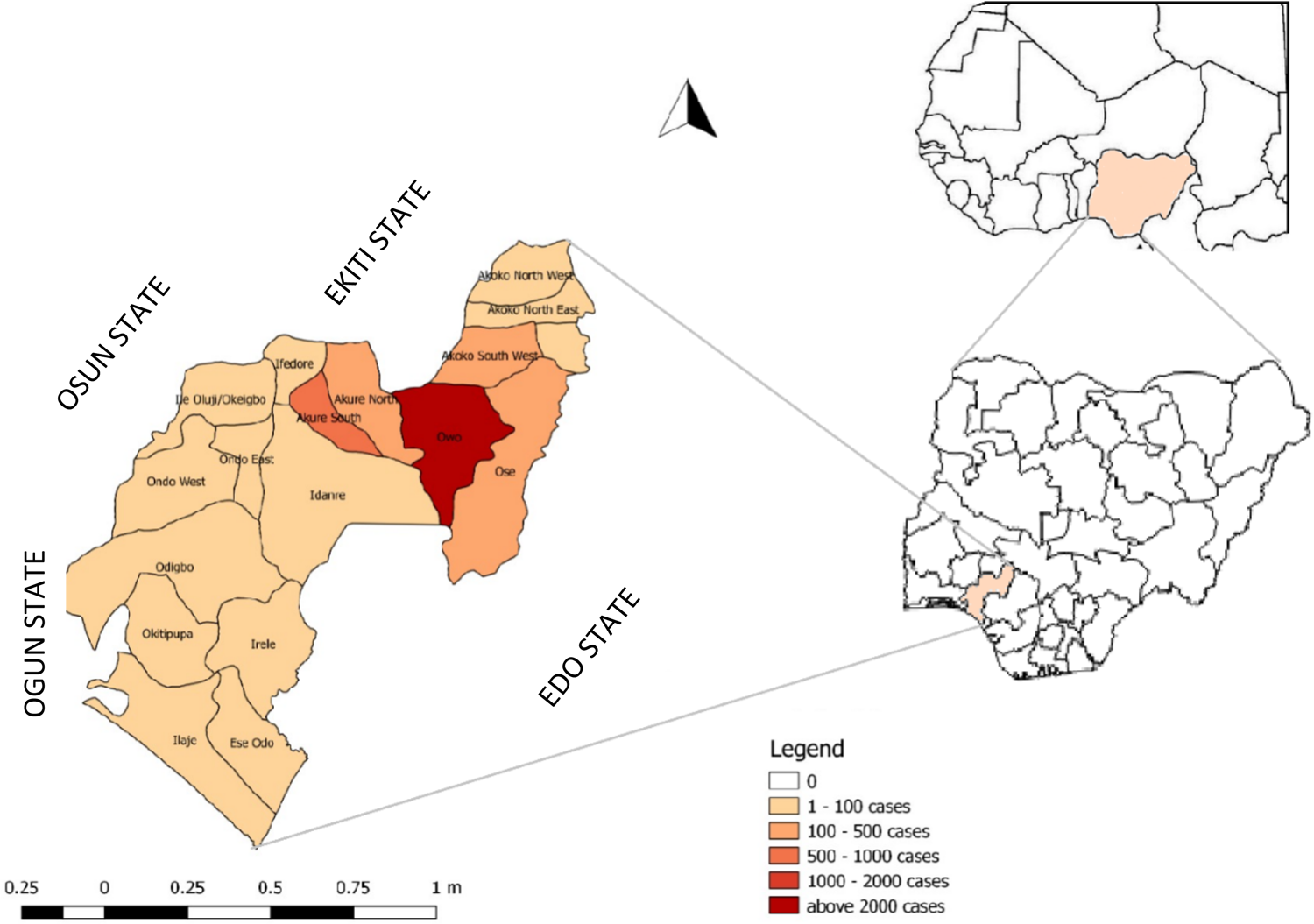
Map of Ondo State showing the 18 Local Government Areas. The intensity of the colour indicates the number of LF cases (inset are Nigeria and West Africa).

**Table 2:**
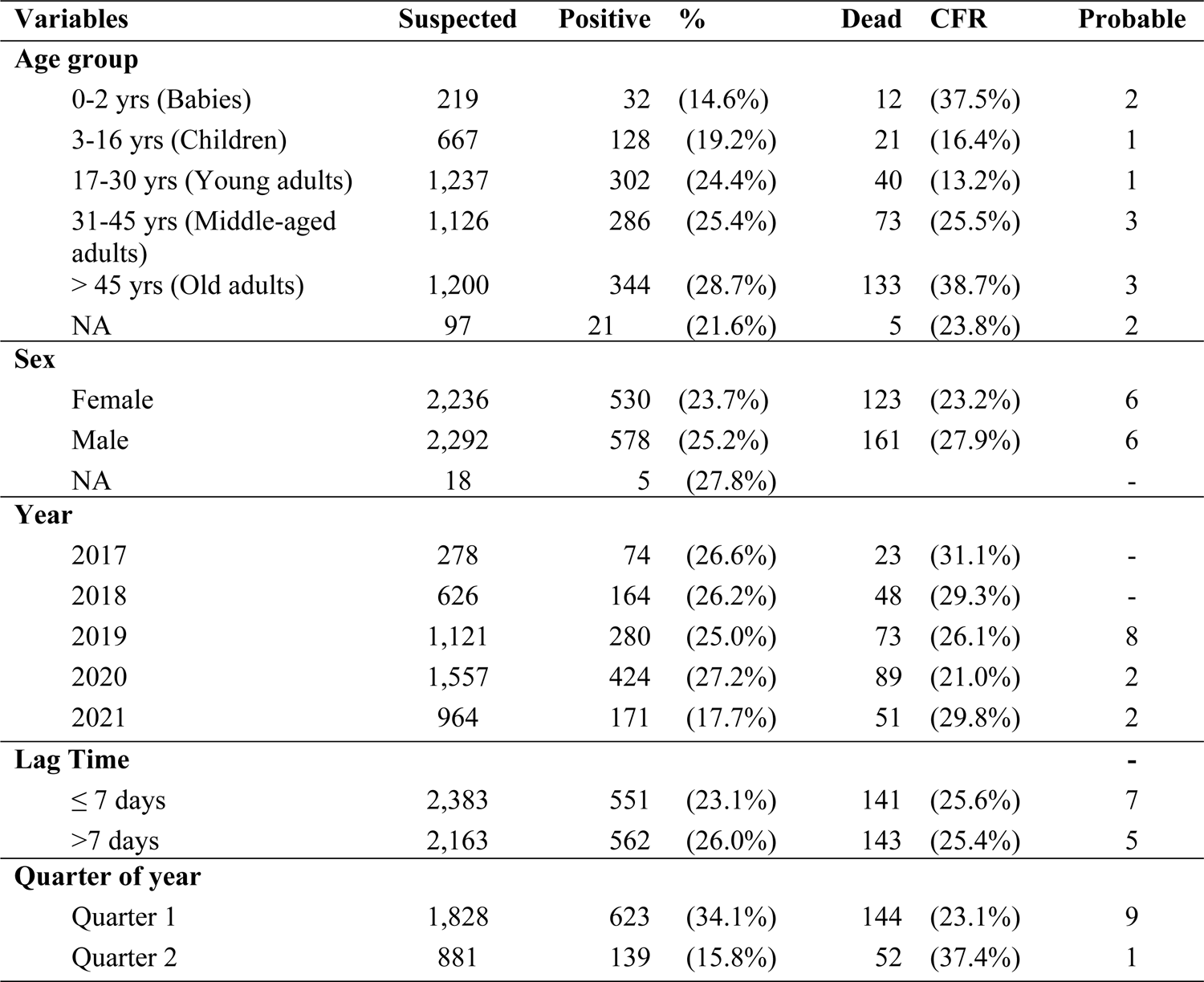
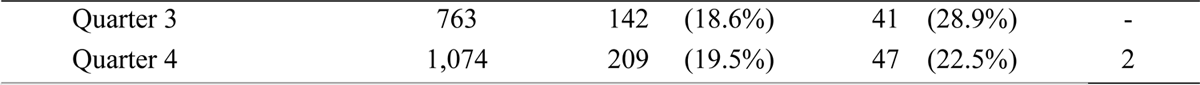
Distribution of suspected and confirmed cases and deaths by selected variables based on Ondo State LF data, 2017-2021.

A total of 1,557 suspected cases were reported in 2020, with 424 (27.2%) confirmed and 89 deaths, giving a CFR of 21.0%. However, 2017 had the worst CFR with 23 deaths from 74 confirmed cases (CFR=31.1%) (Table 2; Figure 4). Majority of the confirmed cases and deaths were reported during the first (623 confirmed, 144 deaths) and fourth (209 confirmed, 47 deaths) quarters of the year. However, most deaths occurred between April and June, the second quarter) (Figure 5).

**Figure 4:**
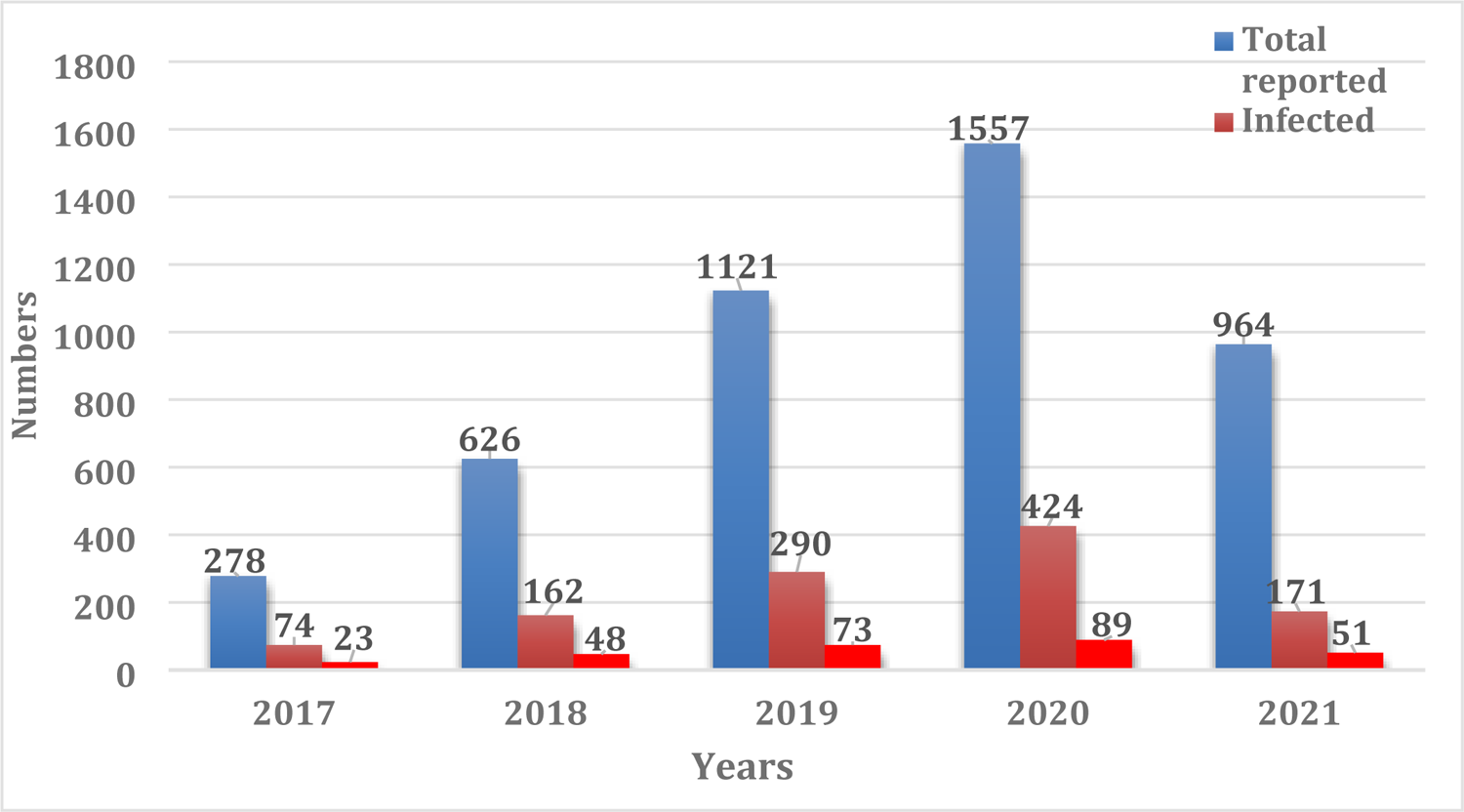
Total number of suspected LF cases, confirmed cases and deaths, Ondo State, 2017 – 2021

**Figure 5:**
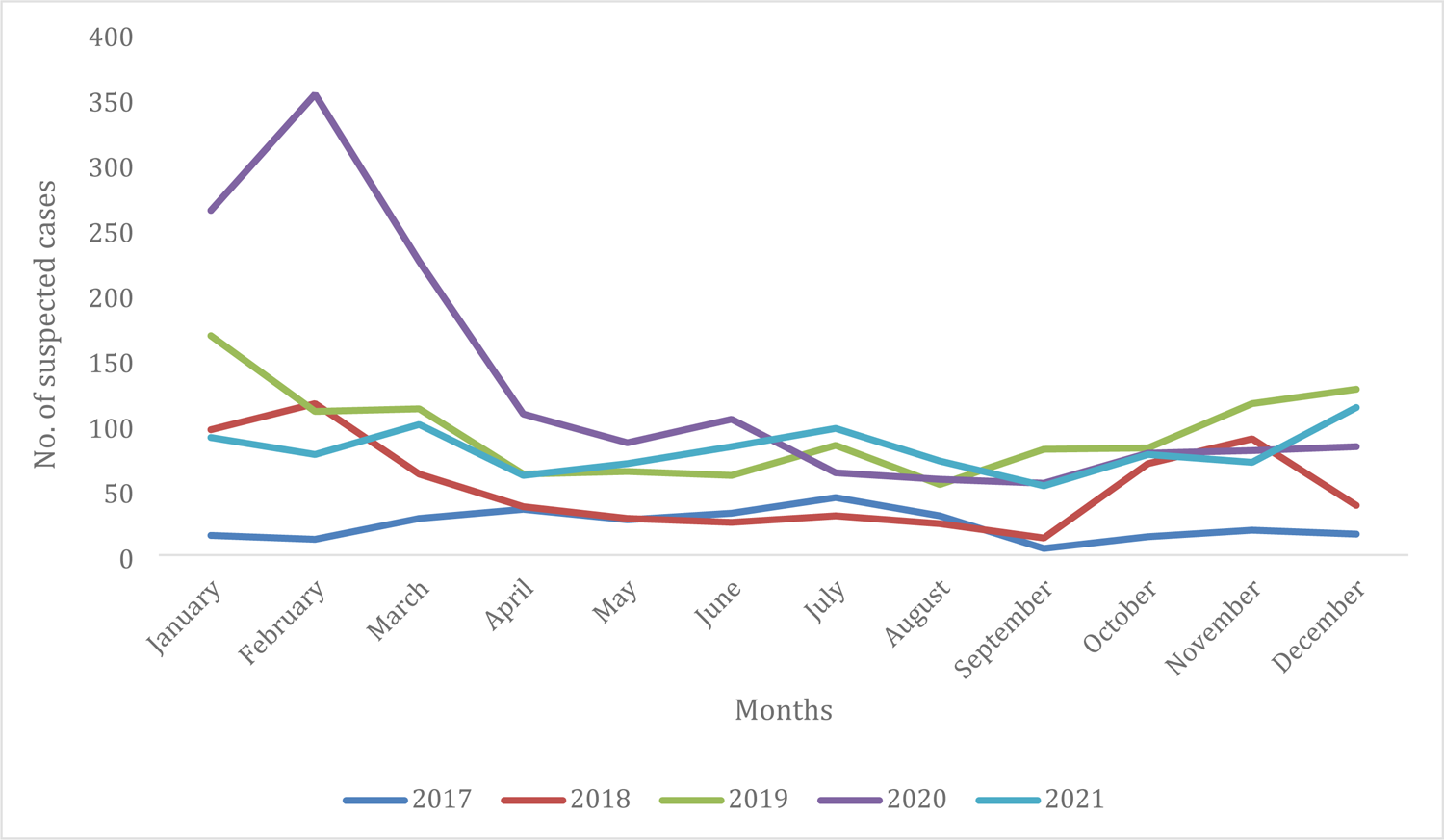
Seasonal trend of suspected LF cases, Ondo State, 2017-2021

The 2021 dataset of 964 infected individuals was further analyzed with the primary objective of estimating the pattern of survival over time after admission to the hospital. (Figure 6A). The data showed that the minimum survival period observed was 5 days, while the maximum survival period after admission to the hospital was 30 days, suggesting a variation in the course of disease in infected patients. The average time to death of LF patients was approximately 17 days (Figure 6A). The survival curve graphically represents the probabilities of survival at different time points after admission to the hospital. The chances of surviving beyond 30 days from the onset of LF are remarkably low. This implies that the disease has a high mortality rate within the first month of infection, making early diagnosis and effective treatment crucial in improving patient outcomes.

**Figure 6A:**
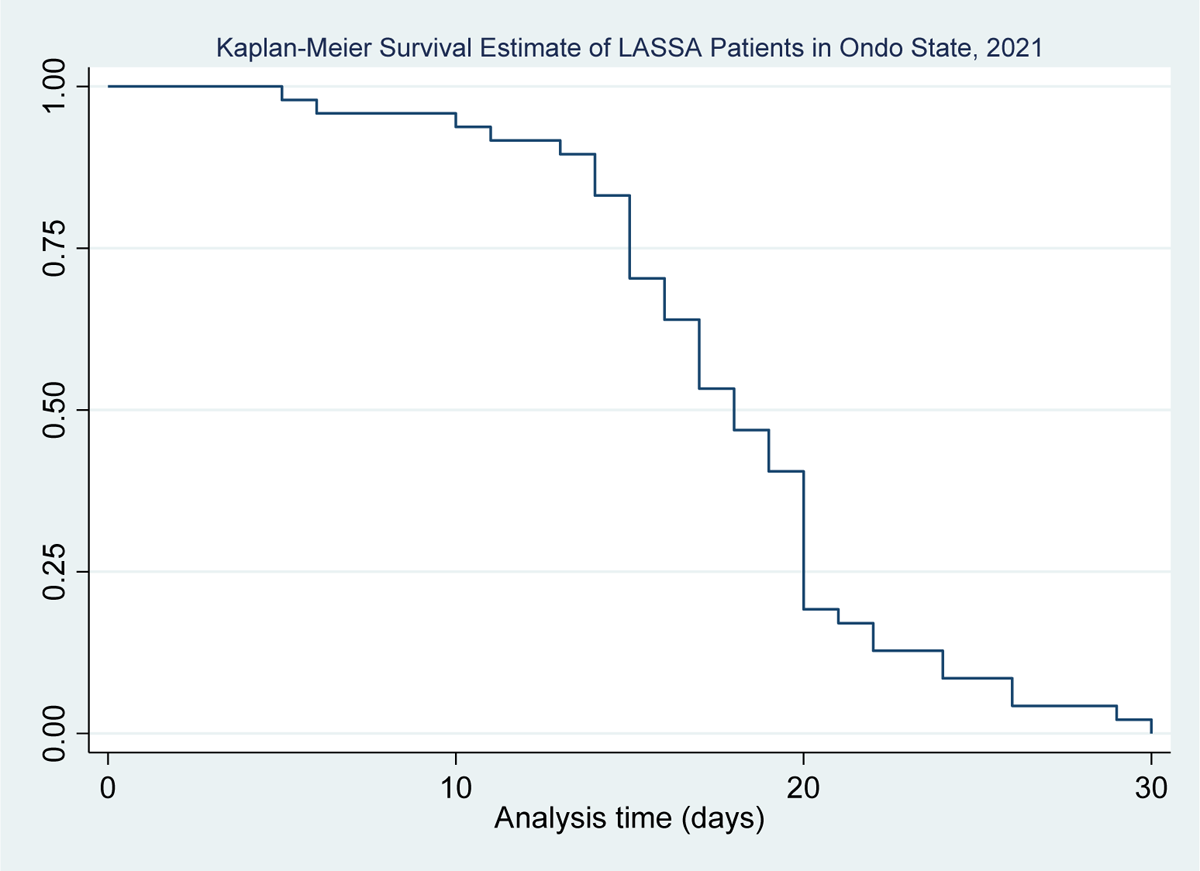
Kaplan-Meier Survival Estimate of LF cases in Ondo State, 2021

In addition to estimating the overall survival function of LF patients, the study conducted a stratified analysis by gender (Figure 6B) to investigate potential differences in survival patterns between male and female patients. The findings from the gender-stratified analysis revealed that male patients demonstrated a lower chance of surviving during the first 10 days of admission to the hospital, suggesting that male patients have a higher risk of mortality in the early stages of infection. However, the analysis also indicated that male patients had relatively improved chances of survival between 10 and 20 days after admission. On the other hand, it was observed that male patients had lower survival probabilities beyond 20 days after admission, suggesting that the disease may have a more severe and prolonged impact on male patients in the later stages of infection. The in-depth analysis of the probability of survival among the LF cases with respect to age is shown in Table 3. The probability of survival is more likely from day 5 to 17, beyond which a significant decrease was observed. This implies that beyond day 17 of infection, there is a significant reduction in the probability of survival of LF patients (Table 3).

**Figure 6B:**
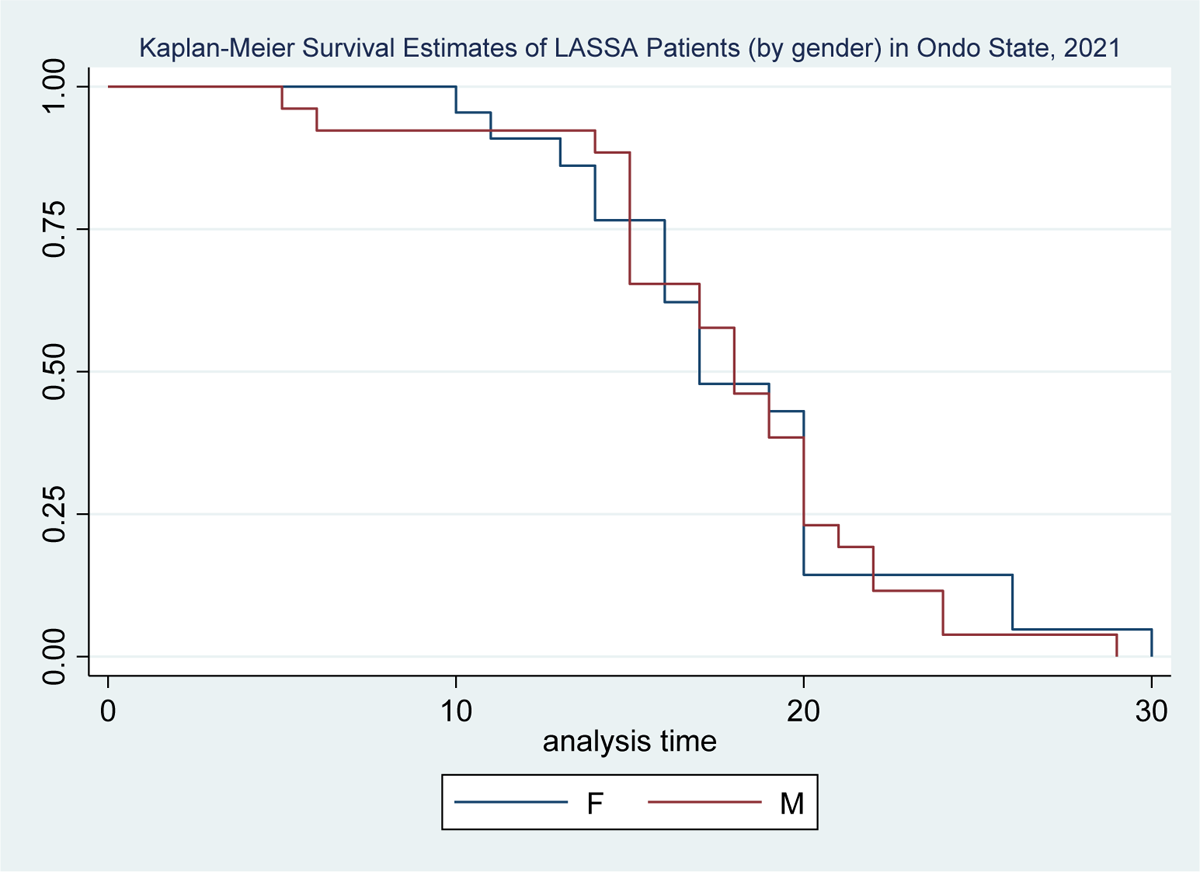
Kaplan-Meier Survival Estimate of LF cases (by gender) in Ondo State, 2021

**Table 3:** Probability of Survival among LF patients in Ondo State (2021)

| Time (days) | Probability of survival | Standard error | 95% confidence interval |
| --- | --- | --- | --- |
| 5 | 0.979 | 0.0206 | 0.861 – 0.997 |
| 10 | 0.937 | 0.0349 | 0.818 – 0.979 |
| 13 | 0.895 | 0.0443 | 0.766 – 0.955 |
| 17 | 0.533 | 0.0728 | 0.381 – 0.663 |
| 21 | 0.171 | 0.0549 | 0.079 – 0.289 |
| 24 | 0.085 | 0.0408 | 0.027 – 0.186 |
| 29 | 0.021 | 0.0211 | 0.002 – 0.098 |

Further analysis of the frequency and distribution of mortality (Table 4) revealed that out of the 171 cases that were identified in 2021, 48 patients (28.1%) died. There were also variations in mortality rates based on age groups. The elderly population, specifically those aged 60 years and above, had a higher proportion of mortality at 8.9%. Similarly, individuals in the 50-59 years age group had a mortality rate of 8.4%. These findings suggest that older individuals had higher risk of succumbing to LF, potentially due to age-related factors such as weakened immune systems or underlying health conditions. Additionally, it was observed that patients from Ose LGA had the highest mortality rate of 11.5%.

**Table 4:** Factors Associated with Mortality among LF Patients in Ondo State, 2021.

| **Result** |  | **Alive** | **Dead** | **p-value** |
| --- | --- | --- | --- | --- |
|  | Negative | 790 (99.9) | 1 (0.1) | **<0.01** |
|  | Positive | 123 (71.9) | 48 (28.1) |  |
| **Age group (yrs)** |  |  |  | **0.012** |
|  | < 20 | 214 (97.7) | 5 (2.3) |  |
|  | 20 – 29 | 156 (98.1) | 3 (1.9) |  |
|  | 30 – 39 | 151 (93.8) | 10 (6.2) |  |
|  | 40 - 49 | 118 (94.4) | 7 (5.6) |  |
|  | 50 – 59 | 87 (91.6) | 8 (8.4) |  |
|  | 60+ | 164 (91.1) | 16 (8.9) |  |
| **LGA** |  |  |  | 0.094 |
|  | Akoko | 73 (93.6) | 5 (6.4) |  |
|  | Akure | 177 (93.7) | 12 (6.3) |  |
|  | Ose | 54 (88.5) | 7 (11.5) |  |
|  | Owo | 576 (96.0) | 24 (4.0) |  |
|  | Others | 33 (91.7) | 3 (8.3) |  |
| **Sex** |  |  |  | 0.388 |
|  | Female | 449 (95.3) | 22 (4.7) |  |
|  | Male | 461 (94.1) | 29 (5.9) |  |

Concerning the distribution of LF over the five-year period, the highest proportion of infected persons (27.3%) was recorded in 2020 (Table 5). This percentage was slightly higher compared to 2017 (26.6%), 2018 (25.9%), 2019 (25.8%), and 2021 (18.0%). Furthermore, over the same period, the analysis revealed that 27.3% of the patients from Owo and Ose LGAs tested positive for LF, making them the areas with the highest proportion of cases. This was followed by 20.7% of cases from Akoko LGA and 19.8% from Akure LGA. In addition, more males (25.5%) than females (23.9%) tested positive for the disease.

**Table 5:** Association between Lassa fever and variables tested.

| Variable | Category | Cases/Infection status |  | OR (95%CI) | P-value |
| --- | --- | --- | --- | --- | --- |
|  |  | Positive | Negative |  |  |
| **Year** | 2017 | 74 (26.6) | 204 (73.4) | 1 |  |
|  | 2018 | 162 (25.9) | 463 (74.1) | 0.96 (0.70 – 1.33) | 0.89 |
|  | 2019 | 290 (25.8) | 833 (74.2) | 0.95 (0.71 – 1.29) | 0.84 |
|  | 2020 | 426 (27.3) | 1132 (72.7) | 1.04 (0.78 – 1.38) | 0.86 |
|  | 2021 | 173 (18.0) | 791 (82.0) | 0.60 (0.44 – 0.82) | 0.00 |
| **LGA** | Owo & Ose | 846 (27.3) | 2257 (72.7) | 1 |  |
|  | Akure | 168 (19.8) | 681 (80.2) | 0.66 (0.55 – 0.79) | 0.00 |
|  | Akoko | 81 (20.7) | 310 (79.3) | 0.69 (0.54 – 0.90) | 0.01 |
|  | Ondo Central | 20 (14.3) | 120 (85.7) | 0.44 (0.28 – 0.72) | 0.00 |
|  | Ondo South | 3 (9.4) | 29 (90.6) | 0.27 (0.08 – 0.91) | 0.04 |
|  | Others | 7 (21.2) | 26 (78.8) | 0.72 (0.31 – 1.66) | 0.56 |
| **Age** | 0-19 | 203 (18.8) | 878 (81.2) | 1 |  |
|  | 20-39 | 432 (24.8) | 1311 (75.2) | 1.43 (1.18 – 1.72) | 0.00 |
|  | 40-59 | 306 (29.6) | 727 (70.4) | 1.82 (1.49 – 2.23) | 0.00 |
|  | ≥ 60 | 184 (26.6) | 507 (73.4) | 1.57 (1.25 – 1.97) | 0.00 |
| **Sex** | Male | 588 (25.5) | 1714 (74.5) | 1 |  |
|  | Female | 537 (23.9) | 1709 (76.1) | 1 (0.87 – 1.15) | 0.97 |

Meanwhile, patients from Akure, Akoko, Ondo Central, and Ondo South were found to be less likely to be infected with LASV compared to those from Owo and Ose LGA. The odds ratios (OR) for the respective locations were as follows: Akure (OR = 0.66; 95% CI = 0.55 – 0.79), Akoko (OR = 0.69; 95% CI = 0.54 – 0.90), Ondo Central (OR = 0.44; 95% CI = 0.28 – 0.72), and Ondo South (OR = 0.27; 95% CI = 0.08 – 0.91). Additionally, patients within the 20-39 years age group were 1.43 times more likely (95% CI = 1.18 – 1.72) to have LF compared to those below 20 years of age. Similarly, patients aged 40-59 years had a higher likelihood (OR = 1.82; 95% CI = 1.49 – 2.23) of being infected with LASV.

In 2017, there were 278 reported cases of LF which resulted in 23 deaths, giving a CFR of 8.3% However, there was a consistent decrease in CFR in the subsequent years, with the rate dropping as low as 5.3% in 2021. Analysis of the CFR in different LGAs of Ondo State showed that Akoko LGA had a CFR of 8.7%, Ondo Central 8.6%, and Akure 6.2%. Furthermore, the CFR varied based on age groups with the highest value of 11.0% observed among patients aged ≥ 60 years, while the lowest CFR of 3.6% was seen among patients below 20 years of age. This highlights the increased vulnerability of older individuals to severe outcomes and complications from LF infection.

The results (Table 6) of regression analysis provided further insights into the factors affecting survival rates. Cases from Akoko LGA were found to be 1.54 times more likely (95% CI = 1.06 – 2.27) not to survive the infection. Also, the results revealed a dose-response relationship between age and the likelihood of not surviving the infection. Patients aged 40-59 years were 2.46 times more likely (95% CI = 1.67 – 3.62) not to survive, while those above 60 years of age had an even higher likelihood (OR = 3.30; 95% CI = 2.22 – 4.92) of not surviving. These indicate that as age increases, the risk of mortality from LF also increases.

**Table 6:** Association between outcome of infection with LASV and variables tested.

| Variable | Category | Infection<br>Dead<br>n (%) | Outcome<br>Alive<br>n (%) | 95% CI | P-value |
| --- | --- | --- | --- | --- | --- |
| **Year** | 2017 | 23 (8.3) | 255 (91.7) | 1 |  |
|  | 2018 | 47 (7.5) | 578 (92.5) | 0.90 (0.53 – 1.52) | 0.79 |
|  | 2019 | 74 (6.6) | 1049 (93.4) | 0.78 (0.48 – 1.27) | 0.39 |
|  | 2020 | 89 (5.7) | 1469 (94.3) | 0.67 (0.42 – 1.08) | 0.13 |
|  | 2021 | 51 (5.3) | 913 (94.7) | 0.62 (0.37 – 1.03) | 0.08 |
| **LGA** | Owo & Ose | 180 (5.8) | 2923 (94.2) | 1 |  |
|  | Akure | 53 (6.2) | 796 (93.8) | 1.08 (0.79 – 1.48) | 0.68 |
|  | Akoko | 34 (8.7) | 357 (91.3) | 1.54 (1.06 – 2.27) | 0.03 |
|  | Ondo Central | 12 (8.6) | 128 (91.4) | 1.52 (0.82 – 2.80) | 0.24 |
|  | Ondo South | 1 (3.1) | 31 (96.9) | 0.52 (0.07 – 3.85) | 0.79 |
|  | Outside the State | 4 (12.1) | 29 (87.9) | 2.24 (0.78 – 6.44) | 0.25 |
| **Age** | 0-19 | 39 (3.6) | 1042 (96.4) |  |  |
|  | 20-39 | 82 (4.7) | 1661 (95.3) | 1.32 (0.89 – 1.95) | 0.19 |
|  | 40-59 | 87 (8.4) | 946 (91.6) | 2.46 (1.67 – 3.62) | 0.00 |
|  | ≥ 60 | 76 (11.0) | 615 (89.0) | 3.30 (2.22 – 4.92) | 0.00 |
| **Gender** | Male | 161 (7.0) | 2141 (93.0) | 1 |  |
|  | Female | 123 (5.5) | 2123 (94.5) | 0.97 (0.69 – 1.37) | 0.93 |
| **Lag**<br>**Period** | 1-7 day | 141 (6.5) | 2036 (93.5) | 1 |  |
|  | 8-14 days | 87 (6.1) | 1339 (93.9) | 0.94 (0.71 – 1.24) | 0.70 |
|  | ≥ 15 days | 56 (5.9) | 889 (94.1) | 0.90 (0.66 – 1.25) | 0.62 |

Furthermore, key informant interviews and environmental assessment carried out identified possible socio-ecological factors that could be fueling the endemicity and outbreak of LF in Owo (the epicenter) and the adjoining towns. The major factors included sun drying of food items by the road side, a local market that serves as a central transaction hub for Owo and the adjoining LGAs as well as the market waste which could attract rats, poor and unsafe sewage disposal, proximity of refuse dump to residential areas and congested urban residential setting with rat infestation (Figure 7).

**Figure 7:**
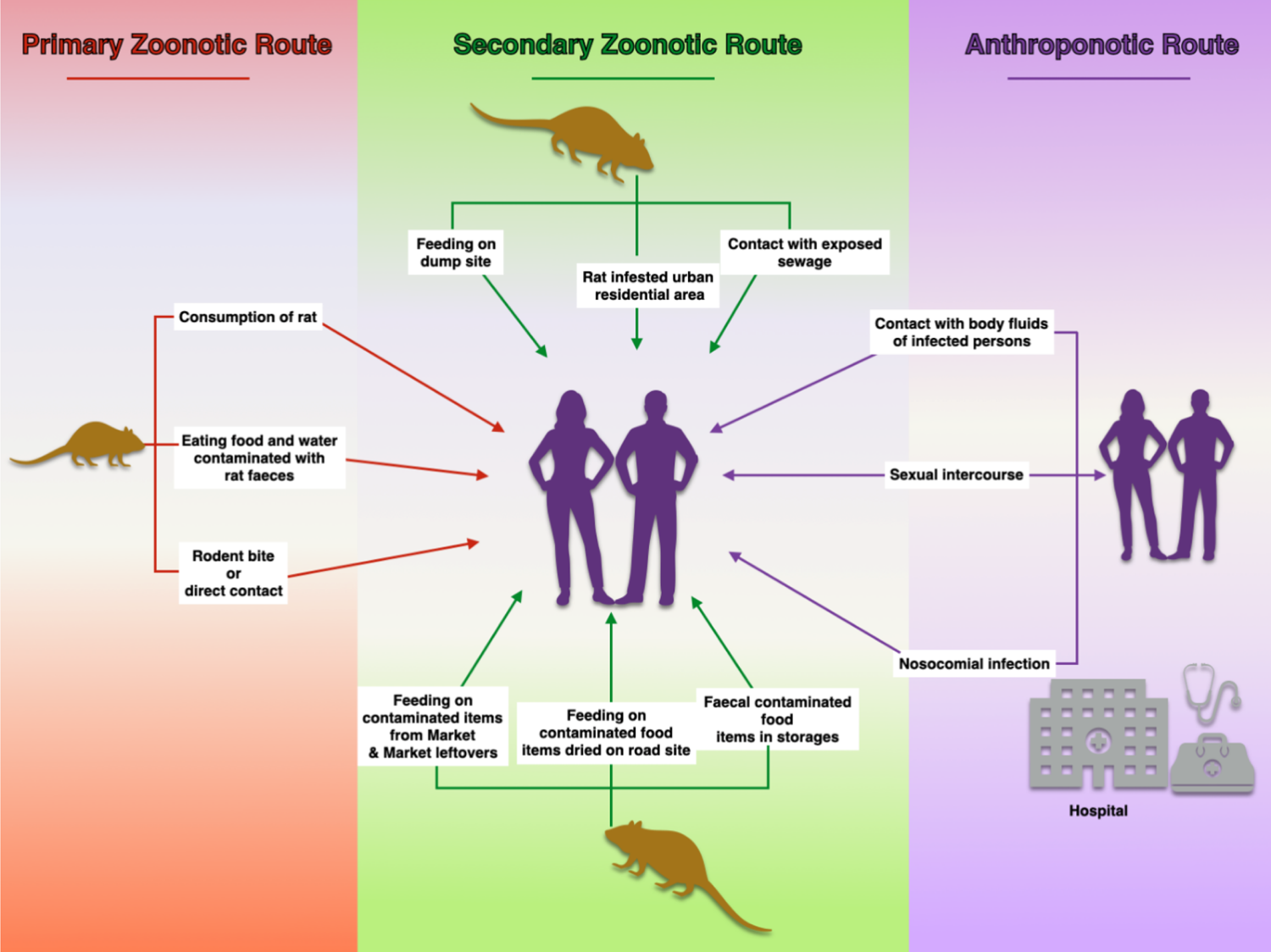
Socio-ecological factors sustaining and exacerbating the endemicity and outbreak of LF in Owo and adjoining towns in Ondo State.

## DISCUSSION

This study involved in-depth epidemiological analysis of a five-year (2017-2021) LF data on Ondo State, southwestern Nigeria following the recent upsurge in reported cases of LF in Nigeria and considering the importance of the study area in the epidemiology of the disease in the country. The findings of the present study revealed a sharp increase in the number of reported cases of LF in Ondo State from 2017 to 2020 and a sharp drop towards 2021. Similar observations have been reported in previous studies conducted across Nigeria [17–19]. One possible reason for this observation is the interventional support of NCDC in 2016 toward improving laboratory capacity for diagnosis of LF as well as the establishment of the Surveillance Outbreak Response Management and Analysis System (SORMAS), an electronic surveillance reporting system in 2018.These interventions have resulted in significant increase in the number of identified and reported cases of LF in the country. Other factors implicated in the increased number of reported LF cases include changing epidemiology which has led to possible redistribution of the rodent reservoirs and identification of hitherto non-conventional hosts and hotspots, human behavioural changes and practices as well as increased number of NCDC-certified testing laboratories, accessibility and transportation of sample material provided by the NCDC laboratories network system [1; 20].

The observed decrease in reported cases in 2021 might not be unconnected with the interruption occasioned by the COVID-19 epidemic of 2020, which redirected the focus of national human and surveillance system resources towards monitoring and keeping-up with the epidemic. The COVID-19 pandemic was a major factor that affected the identification, treatment and management of both infectious and non-communicable diseases globally [21]. For instance, COVID-19 caused a substantial disruption to infectious diseases, like TB, health services and increased vulnerability to diseases because all available resources were diverted towards the response to the pandemic [22]. However, the fact that the situation might be the true reflection of actual case reduction, due to better human awareness, improved behaviour and practices, cannot be ruled out.

Consistent with the findings of previous studies, our result showed peaks of outbreaks between the last quarter and the first quarter of the year [19; 23]. The annual peak between the last and first quarter has been attributed to a combination of factors between the explosion of the *Mastomys* rodents’ population and seasonal coincidence with preparation for planting in which prevailing environmental conditions encourage human-rodent interactions [23; 24; 25]. Lassa fever has evidently been shown to be a climate-sensitive disease whose incidence is associated with seasonal climatic change [20; 26]. Although, the major drivers of reccurring incidence and risk factors for spillover of LF are not well understood, some of the other factors that have been implicated include ineffective food storage, quality of housing, and certain unprofessional agricultural practices such as crop processing and packaging [27].

Our findings showed that a preponderance (28.7%) of the confirmed LF cases during the study period were adults (i.e., 45 years and above). Lassa fever has been previously associated with the productive age of 15-64 years [24; 28]. Importantly, the data from this study specifically identified age as a key risk factor for LF infection, with patients 20 years and above more likely to be infected compared to those who ≤ 19 years. This age range constitutes the active group of the population that are most likely to be exposed to the risk factors of LF such as contact with virus vector and predisposing agricultural practices. It was further observed from our data that higher CFR was obtained for individuals who are 40 years and above, and this age group represents an important risk factor for CFR associated with LF. Individuals who are 40 years and above are more likely to die due to complications resulting from LASV infection. This age group were more exposed to other risk factors of LF, thus the higher CFR. However, it was observed from our data that majority (80.5%) of individuals under consideration were adults (18 years and above), which indicate that most of the people attending the health facilities for LF-related cases were adults. Therefore, this might be the reason age was identified as risk factor for LF and death.

A major observation from our data is that majority (89%) of the cases occurred in Owo and adjoining LGAs (Ose, Akure North, Akure South and Akoko South) of the State. These LGAs share contiguous borders and importantly, two of them share borders with Edo Sate, the epicenter of LF in Nigeria. This is a major public health concern. An important factor in understanding the epidemiology of LF is the interaction that exists between man and his environment [20; 29; 30]. Interestingly, there are three major markets - Ose, Ogbese, and Elegbeda that link the four LGAs. These markets are the main transaction hubs for most market women in these LGAs, within and outside the state, thus serving as a major ecological space for interaction between humans, who are consistently engaged in bushmeat (including rat meat) selling, and the disease vector. Worthy of note is that one of the predominant economic activities among the individuals in these communities (i.e., the LGAs) is granulated cassava (garri) and flour processing as well as selling of bush meat. In addition, one major feature of these markets is that they operate every five days, a practice that is common with peri-urban/rural market settings in Africa. The implication of this is that, apart from the abundance of the risk of exposure, there is also the issue of intensity and frequency of exposure which could serve as a potential factor sustaining and exacerbating the endemicity and outbreak of LF infection in these LGAs of Ondo State. Interestingly, careless food packaging, poor food storage, inappropriate hygienic practices, unsafe waste disposal and poor environmental sanitation have been strongly linked to factors that could encourage the continuous challenges posed by viral diseases like LF [31; 32; 33).

Although, there has not been convincing epidemiological explanation for the reoccurrence of LF outbreaks in Ondo State, other major factors that have been implicated are the issue of spillover and congested urban residential settings that encourage continuous breeding of *Mastomys natalensis* rats (31; 33; 34). There is a need for more in-depth studies to shed light into the fundamental factors that are responsible for the upsurge of seasonal LF outbreaks in Ondo State and neighbouring Edo State. Although a lot of efforts, especially from the NCDC, have been channeled towards early detection, laboratory diagnosis and surveillance of LF, more efforts need to be focused on strengthening local (intra-State) disease surveillance and response implementation into routine laboratory diagnosis and epidemiological surveillance strategy.

Lassa fever is endemic in Ondo State with the epicenter being Owo and the adjoining LGAs. It is important to note that the surveillance data presented here is with a limitation of under-reportage. Therefore, more attention should be focused on scaling up more robust community/social mobilization, novel community enlightenment and health education outreach among the locals in the intrastate epicenters backed with mass media by engaging key community influencers. These will help to prevent, reduce and be better prepared in responding promptly to potential outbreaks at the community healthcare settings and hopefully reduce CFR. Importantly, more resources should be channeled into establishing centralized, easily accessible and functioning LF point-of-care and field diagnostic tools for early detection, such as molecular based rapid diagnostic kit for case confirmation. This should be complemented with reliable and potent drugs as well as vaccines, especially in communities known for outbreaks and associated with abundance of rodent reservoirs. The establishment of strategic, all-encompassing and efficient culturally relevant data and information sharing platform for effective awareness, preparedness and risk communication planning as well as developing local outbreak surge capacities is necessary in reinforcing health systems, emerging disease surveillance and quick response interventions.

Finally, towards community integration and involvement at mitigating the problem of LF at the LGAs and hot-spots, we propose the establishment of Community Action Networks (CANs). This will involve local stakeholders at community level (with equal gender representation) in high-risk settings for Early Warning and Early Response (EWER) using an innovative technology-driven App. Here, a gender-balanced active group of trained like-minded individuals will be set up in disease hot-spots in the state with the sole aim of reporting syndromes of animal diseases to livestock units, syndromes of human diseases (fever and/or body pain, vomiting, jaundice) to dedicated phone lines that will be established. A link with the Integrated Disease Surveillance architecture will be created and existing groups will be adapted for One-Health-sensitive participatory action. The groups will be trained to work with communities and provide support in farming, food storage, pest control, livestock treatment and disease prevention with a focus on community priorities. Ultimately, this will strengthen community level participation and data gathering that will enhance information flow to the existing IDSR system, thus ensuring a sustainable local platform for strengthening surveillance systems for LF within the affected communities in Nigeria. This model that can also be replicated in other LF-endemic West African countries.

## Data Availability

The authors hereby declare the all the data sets used in writing this manuscript will be made available on request at any stage of the manuscript review.

## Acknowledgements

We acknowledge the support of the West Africa One-Health Consortium supported by the International Development Research Centre (IDRC) Canada.

